# Phase I Dose Escalation trials in cancer immunotherapy: Modifying the Bayesian Logistic Regression Model for Cytokine Release Syndrome

**DOI:** 10.1101/2024.06.10.24308712

**Authors:** Matt Chapman-Rounds, Miguel Pereira

## Abstract

We extend Bayesian Logistic Regression to model the dose-toxicity relationship in the setting of phase I dose-escalation/ dose-finding trials for cancer immunotherapies. Immunotherapy drugs are associated with Cytokine Release Syndrome, a systemic immune system reaction that can be mitigated when initial lower doses of the drug are administered to generate immune tolerance. This changes the classic dose-finding problem of determining an optimal safe dose, to a more complex problem where the search is for both the optimal safe dose and the dose regimen that allows patients to quickly and safely reach that dose without CRS. As part of solving this methodological challenge, we show how to jointly model CRS and non-CRS toxicities, which have distinct mechanisms, while controlling for the overall toxicity rate to make dose-escalation decisions.

## 1 Introduction

The goals of a phase I, first-in-human, dose-finding clinical trial are to collect information on the safety and tolerability of a new drug, identify the maximum tolerated dose (MTD), and identify one or more doses to assess in a phase II study (recommend phase II doses or RP2D)[5].

The MTD is the highest dose which can be administered to humans which best balances toxicity (i.e. probability that a subject experiences a serious adverse event, also called a dose-limiting toxicity or DLT). Generally, both drug efficacy and drug toxicity increase with dose. In order to find the MTD, the general approach of a phase I trial is to start at a low dose and carefully escalate to higher doses in subsequent groups of participants (usually size 3-6). As the dose increases, DLTs will start to occur, and at some point we will stop increasing the dose when we conclude that the toxicity of any further increase will be too high.

The use of Bayesian response adaptive models for phase 1 studies has been advocated by the European Medicines Agency adopted guideline on small populations [3] and by Rogatko and colleagues [1, 7] and was one of the key elements of the FDA’s Critical Path Initiative.

The Bayesian Logistic Regression Model (BLRM) using Escalation with Overdose Control (EWOC) is an increasingly common methodology used to carry out dose escalation [1]. It performs significantly better than the widely used 3+3 design, showing better targeting rates (30% in the 3+3 vs. 60% in the BLRM) with fewer patients treated above the MTD. Additionally, the BLRM considers the previously observed data and provides added flexibility as it allows cohorts of different sizes and escalation to intermediate dose levels [6].

The advent of novel treatments in oncology has lead to the development of multiple immuno-therapeutic agents whose action is to stimulate the immune system to act against the tumour cells. An import toxicity caused by these drugs is Cytokine Release Syndrome (CRS), a systemic inflammatory reaction that occurs when many immune cells are activated and release large amount of cytokines into the body[4].

CRS is more likely to occur at lower doses in naïve patients vs. patients whose immune system has already been primed to the drug. To avoid CRS therefore, subjects are first exposed to lower doses of a drug compound in preparation to receiving the target dose. As such, modelling dose-toxicity in this setting poses three novel problems:

1. Instead of modelling a single dose level, we need to model a dose *regimen* whereby patients receive several lower doses of the drug before reaching the target dose. The goal is now to find the maximum tolerated dose regimen (MTD-R), which is a much more challenging problem than finding the MTD because we need to consider multiple additional factors including the number of prior doses, and the shape of the dosing curve.
2. To capture the ameliorating effects of exposing subject to preparatory lower doses, we need to model both CRS toxicities and non-CRS toxicities, and then combine these models to estimate the joint posterior probability of toxicity.
3. In case of toxicity, we need to identify the step(s) in the dose regimen that need(s) adjustment, to better drive dose regimen-escalation decisions during the phase I trial.

This work focusses on extending the BLRM to accommodate modelling of both CRS and non-CRS toxicities in dose regimen-finding phase I trials for immunotherapy drugs. Our approach builds on an adaptation of the methodology proposed by Gerard et al. [2].

## 2 Models

The standard BLRM defines the probability of DLT, *P*(*DLT*) at dose *D* as:

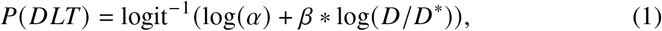

Where *α* and *β* are the parameters of the model and *D*^*\**^ is the reference dose such that the odds of DLT at dose *D*^*\**^ are equal to *α*.

At each stage in a standard phase I trial, the posterior distributions of parameters *α* and *β* would be updated via Bayes’ Rule based on the incidence of DLTs observed in subjects thus far. These posteriors would then be used to generate the probability of *P*(*DLT*), i.e. the posterior probability density over possible true DLT rates at the next dose, or doses, under consideration. Doses can then be described as being in the underdose, overdose, or target dose range. Using thresholds often used in Oncology trials, these ranges are as follows:

- **Underdose:** Probability *P*(*DLT*) < 0.16 is greater than 0.5, and not
- **Target dose:** Probability 0.16 <= *P*(*DLT*) < 0.33 is greater than 0.5, and not
- **Overdose:** Probability *P*(*DLT*) >= 0.33 is greater than 0.25

If the current dose satisfies the requirements for target dose, and the next possible dose is an overdose, then escalation stops and the dose is declared the MTD. If the next dose is an overdose, but the current dose does not satisfy the condition to be the target dose, the dose might still be considered as the RP2D. Alternatively, escalation to a slightly higher, intermediate dose level may be tested to accurately ascertain the MTD.

### 2.1 Modified BLRM

In the context of CRS, to find the maximum tolerable dose *regimen* (MTD-R), rather than modelling simply the relationship between dose and response, we need to model the relationship between a vector of doses, and response.

Our approach is to track two distinct types of adverse events - those due to CRS, and those which are due to other types of toxicity (non-CRS adverse events). The probabilities of each type of event are modelled separately, and assumed to be independent.

Consider a dose regimen which includes an escalation period and a steady state period. For example, a four-step regimen might include the doses 5, 10, 25, 50, 50, 50 at intervals of several days to complete a cycle whose ‘steady state’ or target dose dose is 50*μ*g. We call the period between two doses a ‘stage’; for example, the above is a four-step dose regimen (four distinct doses), but has 6 stages (the final stage will extend from the final dose until end of the safety follow-up).

As with the standard approach, the goal is to generate the posterior probability of *P*(*DLT*) for any dose regimen, and determine if this regimen corresponds to a target dose, underdose, or overdose. Figure 1 plots example distributions after 0 out of 3 subjects experienced a DLT when given the first regimen, and 1 out of 4 subjects experienced a DLT when given the second dosing regimen. Regimens are indexed by their steady state dose; the third and next regimen to be considered in Figure 1 is the four-step regimen described above, with a steady state dose of 50*μ*g.

**Fig. 1.**
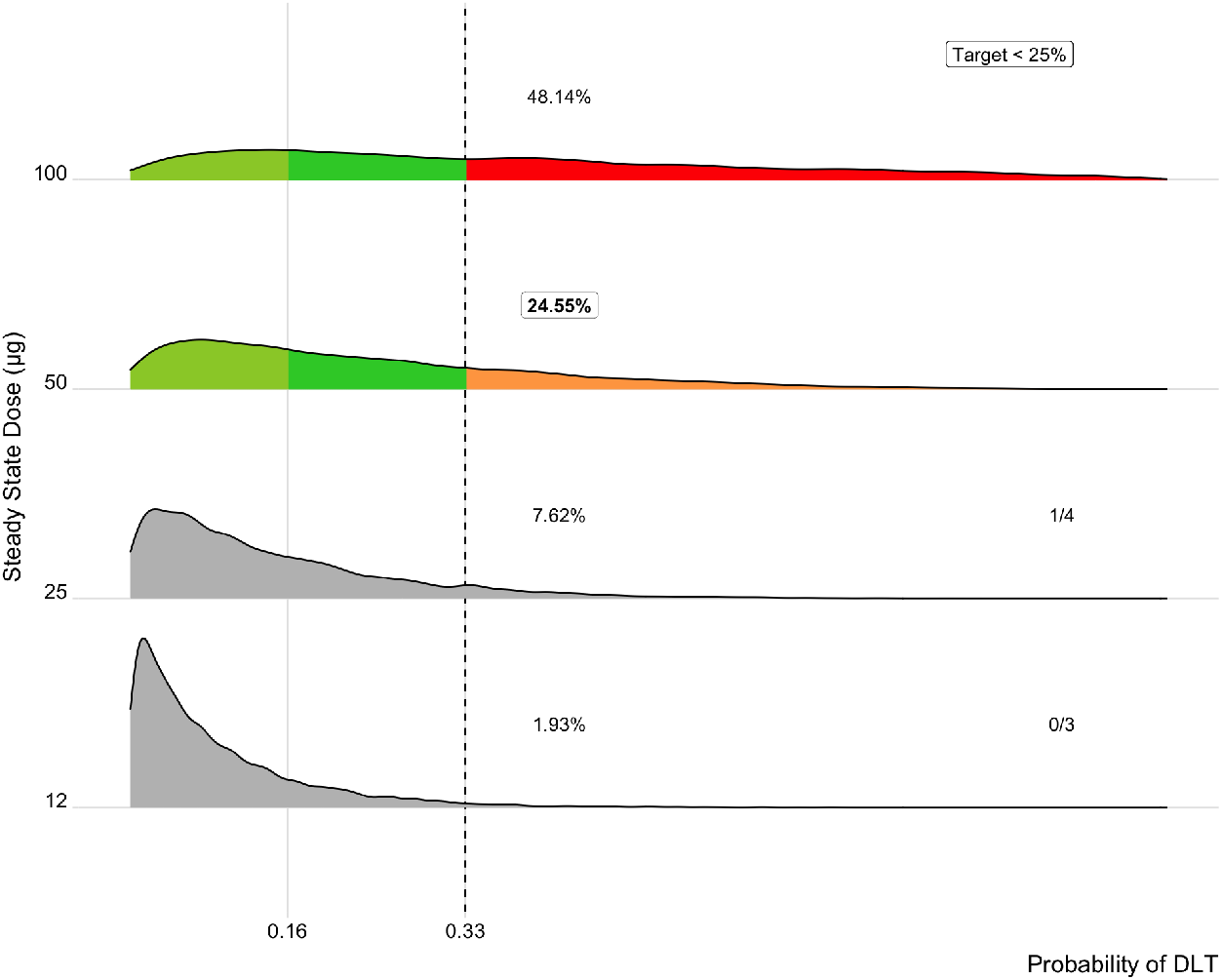
Posterior densities after 1 DLT observed out of 4 subjects in the second regimen, and 0 DLTs observed out of 3 subjects in the first regimen. Red colour indicates that the probability mass >= 0.33 is > 0.25.

#### 2.1.1 CRS Model

We assume that for each stage *i*, the principle positive correlate with probability of CRS in stage *i* is *C*_*max*,*i*_, the maximum concentration reached after dose administration. Whilst we lack the pharmacokinetic data to capture this directly (this data is seldom available at the time of dose escalation), we will take the approach of approximating it using the dose at the start of stage *i, D*_*i*_, i.e.

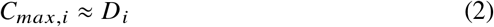

This is counter-balanced by the exposure to the drug prior to stage *i* (since exposure decreases the likelihood of CRS). We approximate this exposure using the ‘area under the dose’, AUD_*i*_,

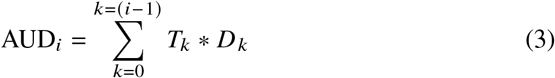

where *T*_*k*_ is the length of stage *k*.

For any stage *i* we define the probability that a CRS-related DLT occurs within that stage, *P*(CRS_*i*_) as

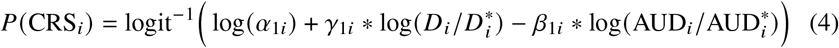

Where 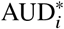 is the AUD for the *i*th stage of a selected reference regimen, and 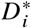 is the dose for the *i*th stage of that reference regimen. Note that it is possible to choose references from different dose regimens - however, for simplicity, we will assume here that a single dose regimen is chosen as reference, and used as such across all stages. If this assumption is not kept, then *α* no longer has a particularly informative interpretation.

The probability that no CRS occurs in a subject is the joint probability of CRS not occuring in each stage, which assuming *S* stages, we can write as *P*(no CRS_1_, …, no CRS_*i*_, …, no CRS_*S*_).

Firstly, we note that *P*(no CRS_*i*_) is independent of all *P*(no CRS_*j*_), *j* > *i*. This follows from the direction of causality.

Secondly, we note that *P*(no CRS_*i*_) ⊥ *P*(no CRS_(*i*−1)_ |AUD_*i*_), i.e. that the probability of CRS/no CRS for stage *i* is conditionally independent of the probability of CRS/no CRS for the previous stage (*i* − 1) given the dose exposure up until the end of that previous stage. This follows from the fact that our model’s calculation of AUD_*i*_ includes all components contributing to the probability of CRS at the previous stage (*D* _(*i*−1)_ and AUD_(*i*−1)_). Given this Markov property for dose stages, it follows that *P*(no CRS_*i*_) **⊥** *P*(no CRS_*j*_ |AUD_*i*_), for all *j* < *i*.

Hence we can write

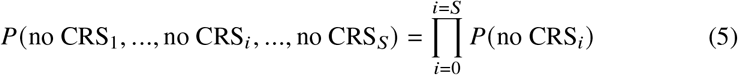

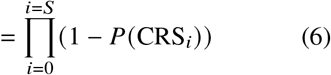

which is the probability that no CRS occurs. As CRS either occurs or does not occur, the probability that CRS occurs is simply

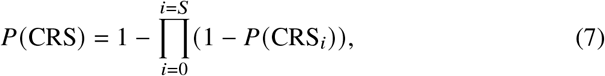

which completes our model of CRS related DLTs.

#### 2.1.2 Non-CRS Model

This model is similar to the standard BLRM, with the exception that it uses uses the cumulative dose over the regimen, *D*_*cum*_, to predict the probability that any other DLT occurs. Note that this model does not consider where DLTs fall in terms of stages, but rather estimates the probability of a non-CRS DLT, *P*(non-CRS), occuring at any time. We write

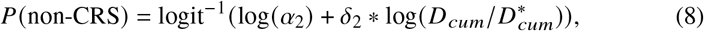

which completes our model of non-CRS related DLTs.

#### 2.1.3 Joint Model

The probability of any DLT occuring is the probability that either a CRS or non-CRS DLT occurs. This is 1 minus the probability that neither a CRS or non-CRS DLT occurs. We assume that CRS and non-CRS DLTs, as modelled, are independent. Hence

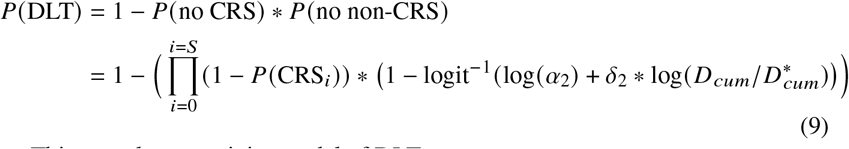

This completes our joint model of DLTs.

### 2.2 Prior Elicitation

In the context of Bayesian analysis, it is necessary to specify priors. As a phase I trial generally marks the first introduction of the study drug to humans, there is little information about the dose-tolerance relationship for CRS (characterised by parameters [*α*_1*i*_], [*β*_1*i*_], [*γ*_1*i*_], *i* ∈ {1, …, *S*}) and non-CRS DLTs (parameters *α*_2_, *δ*_2_). The uncertainty about this relationship is therefore specified as a set of minimally informative priors.

The priors are defined such that for the prospective dose regimens defined at the start of the trial we expect to have minimal toxicity at the lowest dose regimen, and some toxicity (e.g. *P*(*DLT*) ≈ 25%) at the highest regimen.

The prospective dose regimens are selected by pharmacokinetic and clinical experts based on available data from animal studies and other sources of information such that the first dose regimen should be safe, but by the highest regimen we should have encountered some DLTs. If no DLTs have been encountered by the end of the study, but efficacy is still increasing with dose, then this indicates that too many subjects have been given low-efficacy doses, and the escalation should have been more rapid.

Setting our priors such that we expect some toxicity in the higher regimens reflects our belief that the dose regimens have been correctly chosen such that we will escalate from a safe dose to a dose which begins to produce DLTs.

#### 2.2.1 Standard BLRM

In the context of the standard BLRM, for a range of prospective doses from *D*_1_ to *D* _*N*_, we might assume that *P*(*DLT* |*D*_1_) = 0.01 and *P*(*DLT* |*D* _*N*_) = 0.25. The equation for *P*(*DLT*) for the standard BLRM only has two parameters *α* and *β* (see Equation 1); assuming *α* and *β* are Gaussian distributed we can use these two assumed toxicities to solve for point estimates for prior means *μ*_*α*_ and *μ*_*β*_. Setting the vector of standard deviations [*σ*_*α*_, *σ*_*β*_] to reasonably large values, e.g. [2, 1] gives us a first estimate of the prior.

Following this step, the behaviour of the BLRM with this initial prior estimate is checked by simulation. Several basic scenarios which might be examined include:

- If no DLTs have been observed to date, the BLRM should always recommend to escalate (i.e. the next dose under consideration should be estimated to be either underdose or target dose by the BLRM).
- If 3/3 DLTs are observed at any dose, the BLRM should recommend to de-escalate the dose (i.e. the current dose and the next dose under consideration should be estimated to be overdoses), possibly by more than one dose level.
- If 1/3 DLTs or 2/3 DLTs are observed at any dose, the BLRM should recommend either expansion or de-escalation. Which is appropriate would depend on several factors such as the relative size of the jump from the previous dose, and would be subject to the judgment of the expert eliciting the prior.

If the required behaviour is not met, then the prior can be adjusted by changing the assumed true probability of DLT at *D* _*N*_, and re-solving.

#### 2.2.2 Extended BLRM

Rather than two parameters, the extended BLRM has (3 *\* S*) + 2 parameters. As such, assuming low toxicity at the starting dose regimen and high toxicity at a final or high dose regimen will not allow us to estimate the means of the priors for each parameter (as the matrix of the two equations generated by this assumption will be non-invertable).

To handle this, we first treat the probability of CRS and non-CRS DLTs separately. This has the advantage of allowing us some flexibility in the assumed toxicities (for example, we might expect the probability of DLTs due to non-CRS related causes to be higher than that of CRS DLTs). As non-CRS DLTs are modelled by only two parameters, this gives us an initial estimate of the prior for the non-CRS DLT BLRM.

To gain an initial estimate of the prior for the CRS DLT BLRM, our approach is to assume that the CRS model has the form 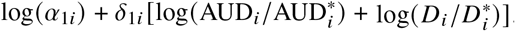. We assume that toxicity at the final dose regimen decreases linearly with stage, and solve the two-point problem for *α* and *δ* for each stage. We then set *β*_1*i*_ = *δ*_1*i*_ − *ϵ M*_*aud*_, and *γ*_1*i*_ = *δ*_1*i*_ + *ϵ M*_*D*_, where *ϵ* is some constant (here 0.01), and *M*_*\*\**_ is a scaling factor which adjusts *ϵ* roughly by the relative magnitude of mean AUD to mean D across all regimens. Again, we assume Gaussian priors with these point estimates as the means, and wide standard deviations.

Once the initial prior estimates have been established, the priors are adjusted until the model exhibits satisfactory predicted behaviour as with the standard BLRM.

This approach works well in practice, but is currently an open area of research; finding a prior which is well behaved is something of an artform: the above approach to find point estimates of the parameter prior is extremely heuristic, and relies on the fact that the priors are kept minimally informative by the addition of noise for its validity. Mixture priors may often be used if there are competing sources of information, but this adds a further layer of complexity beyond the scope of this work.

## 3 Methods

To demonstrate the improved efficacy of the modified BLRM in the setting where dose regimens are being used in place of single doses, compared with the standard BRLM, we simulate the operating characteristics of the models across three scenarios:

1. **Priors Correct** In this scenario the priors of the BLRM are correct. The number of DLTs in a simulated trial are generated by drawing a sample from the prior. This simulates a scenario where we have elicited priors which reflect the true dose-tolerance relationship.
2. **2. High Toxicity** The priors of the BLRM are an under-estimate. The number of DLTs in a simulated trial are generated with probability 125% that of the prior. This is a scenario where we have elicited priors which consistently under-estimate the probability of toxicity; the BLRM should swiftly discard its priors in favour of evidence of high toxicity.
3. **3. Low-High Toxicity** The priors of the BLRM are correct for the first four regimens under consideration. However, in the last three regimens the toxicity spikes sharply: true probability of DLT for trials simulated for these regimens are 25% higher than predicted by the prior. This simulates a worst case scenario where the BLRM has both prior and evidence from lower dose regimens supporting a particular dose-tolerance relationship, but this does not generalise correctly to higher dose regimens. The BLRM should quickly adjust its posteriors to reflect the new evidence at higher doses.

Each scenario assumes that we have pre-selected 7 regimens with increasing dose. Each regimen has four distinct doses (‘steps’) across five stages. Table 1 outlines the prospective dose regimens used for simulation.

**Table 1.** Prospective Dose Regimens for simulation. ‘DR’ = Dose. All values in *μ*g.

|  | Day 1 | Day 5 | Day 10 | Day 14 | Day 21 |
| --- | --- | --- | --- | --- | --- |
| **DR1** | 1 | 2 | 5 | 15 | 15 |
| **DR2** | 3 | 6 | 20 | 75 | 75 |
| **DR3** | 10 | 20 | 50 | 250 | 250 |
| **DR4** | 25 | 50 | 150 | 550 | 550 |
| **DR5** | 25 | 50 | 300 | 1000 | 1000 |
| **DR6** | 25 | 50 | 300 | 1500 | 1500 |
| **DR7** | 25 | 50 | 300 | 2850 | 2850 |

For each scenario, 500 trials were simulated as follows, where *DR*_*C*_ denotes the current regimen under consideration:

For each dose regimen *DR* _*j*_, for each scenario, RJAGS was used to draw 10000 samples of *P*(non-CRS|*DR*_*j*_) using Equation (8) and Gaussian priors with *μ*_*α*2_, *μ* _*δ*2_, *σ*_*α*2_, *σ*_*δ*2_ generated assuming *P*(non-CRS|*DR*_1_) = 0.05 and *P*(non-CRS|*DR*_7_) = 0.2.

### Algorithm 1

Simulation algorithm for a single trial

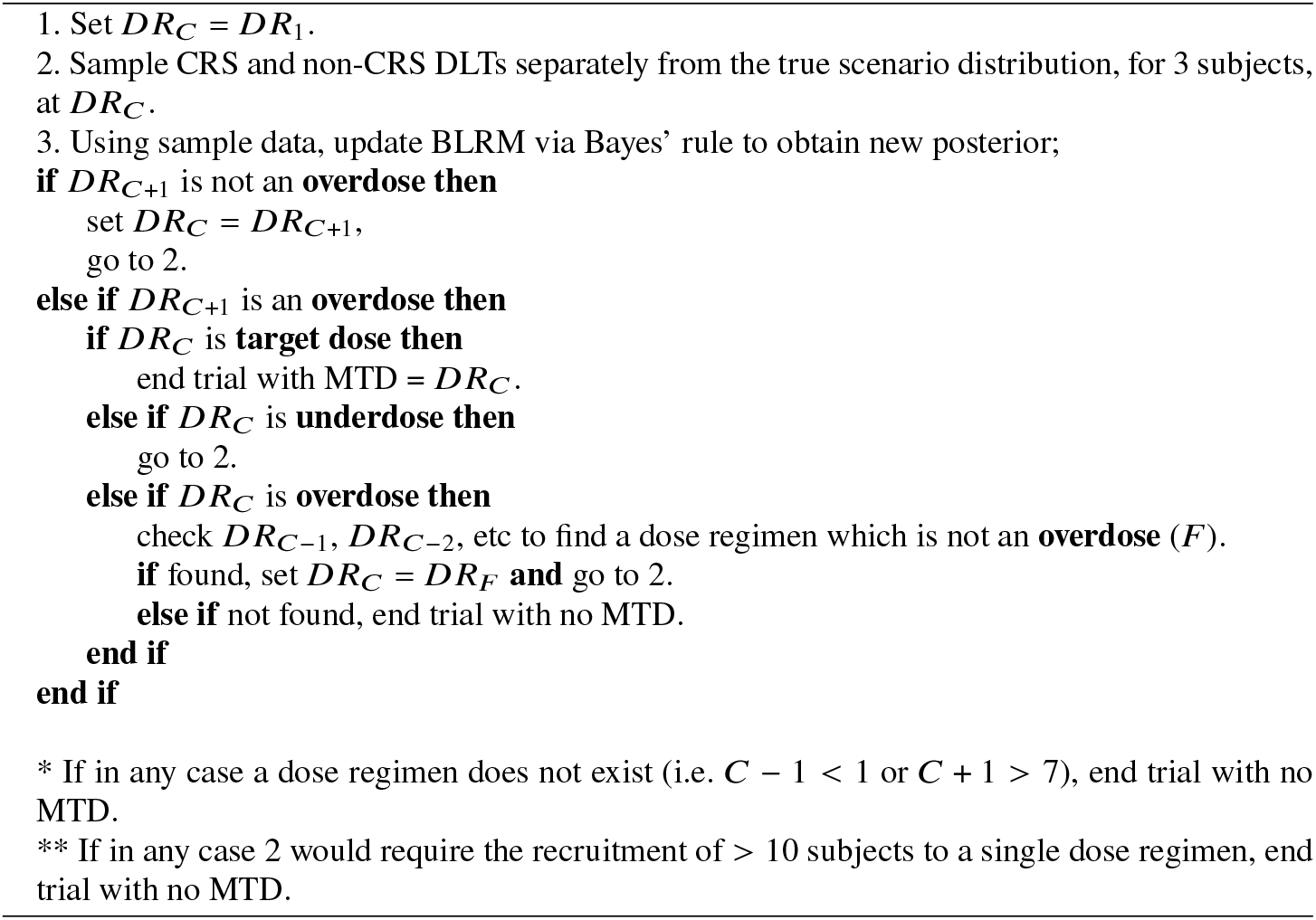

For each stage *i*, for each dose regimen *DR* _*j*_, for each scenario, 10000 samples were drawn from *P*(CRS_*i*_ |*DR* _*j*_) using Equation (7) and Gaussian priors on [*α*_1*i*_], [*β*_1*i*_], [*γ*_1*i*_], *i* ∈ {1, …, *S*} generated assuming *P*(CRS|*DR*_4_) = 0.1 and *P*(CRS|*DR*_7_) = 0.3 with *ϵ* set to 0.01.

To simulate whether, for a single patient, a CRS DLT occurred at a specific stage or a non-CRS DLT occurred at any point during a regimen’s administration, a single sample of *P*(non-CRS|*DR*_*j*_) or *P*(CRS_*i*_ |*DR* _*j*_) was drawn uniformly at random. If the sample was >= 0.5, DLT occurred, if < 0.5, no DLT occurred. If a CRS DLT occurs at an early stage (i.e. *i* < 5), the subject is assumed to have withdrawn from the study, and is not counted for later stages when updating the posterior.

The standard BLRM was simulated using both the steady state dose from each regimen *j* as *D* _*j*_, and the cumulative dose across each regimen *j* as *D* _*j*_. These gave very similar results: for clarity only the results from the cumulative dose approach are given.

For the priors to match between the standard BLRM and the extended BLRM, the extended BLRM prior defined above was used to estimate the overall probability of DLT at DR1 and DR7 (using Equation 9), and then these estimates were used to generate point estimates of the means of the priors on *α* and *β* for the standard BLRM, as described in Section 2.2.1.

## 4. Results

Table 2 shows the operating characteristics for each of the three scenarios described, for both the standard BLRM and the modified BLRM. These are the percentage of trials which either stop with no MTD declared, or which declare an MTD with the true DLT rate for that dose regimen in either the overdose (0.33-1], target dose (0.16-0.33], or underdose [0-0.16] ranges.

**Table 2.**
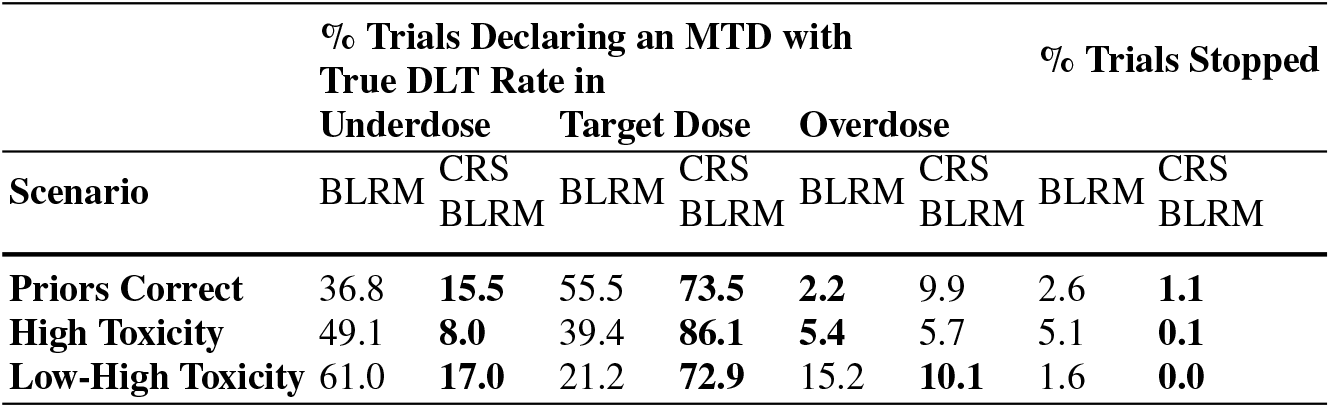
Operating Characteristics of standard BLRM vs CRS BLRM in three scenarios: where the prior beliefs about toxicity are correct, where the toxicity is higher than expected, and where the toxicity is as expected for the first few doses, but then increases much more rapidly than predicted.

In all three scenarios, the modified BLRM correctly finds the target dose more often than the standard BLRM - a predicted success rate of over 70% in each case. The modified BLRM also declares an underdose as MTD less often than the standard BLRM in each scenario.

However the modified BLRM declares an overly toxic dose MTD more often than the standard BLRM both when the priors are correct and in the high toxicity scenario. It is less easily tricked than the standard BLRM by the low-high scenario however: the standard BLRM struggles in this setting as it tends to either declare MTD in a toxic setting or de-escalate aggressively and declare MTD with true DLT rate in the underdose range.

Overall, these results show that the modified BLRM is better suited than the standard BLRM to modelling the nuance of a setting where we have to escalate between dose regimens.

### 4.1 Real World Use

In a real phase I trial, the BLRM will be used to inform the decisions of a dose-escalation committee (DEC). This is a decision-making body which meets after each dose regimen has been tested in a small cohort of subjects to determine whether the dose regimen should be escalated, expanded, de-escalated, or whether the trial should discontinued due to safety concerns.

Both the classic BLRM and our modified variant can be updated at each escalation step of the trial, given the data observed, to provide a full posterior over the dose-response relationship. This means that they can provide estimates of both the probability of overdose at the next potential dose level, and any intermediate dose level, allowing the DEC flexibility in its decisions.

One additional advantage of the modified BLRM is that it allows us to dig more deeply into this posterior: we can split by CRS DLTs versus non-CRS DLTs, and examine the effect of changing specific parts of the regime on the total posterior.

Figure 2 shows the contribution to the total posterior for the third regimen from each pre-steady state regimen stage, for CRS DLTs specifically.

**Fig. 2.**
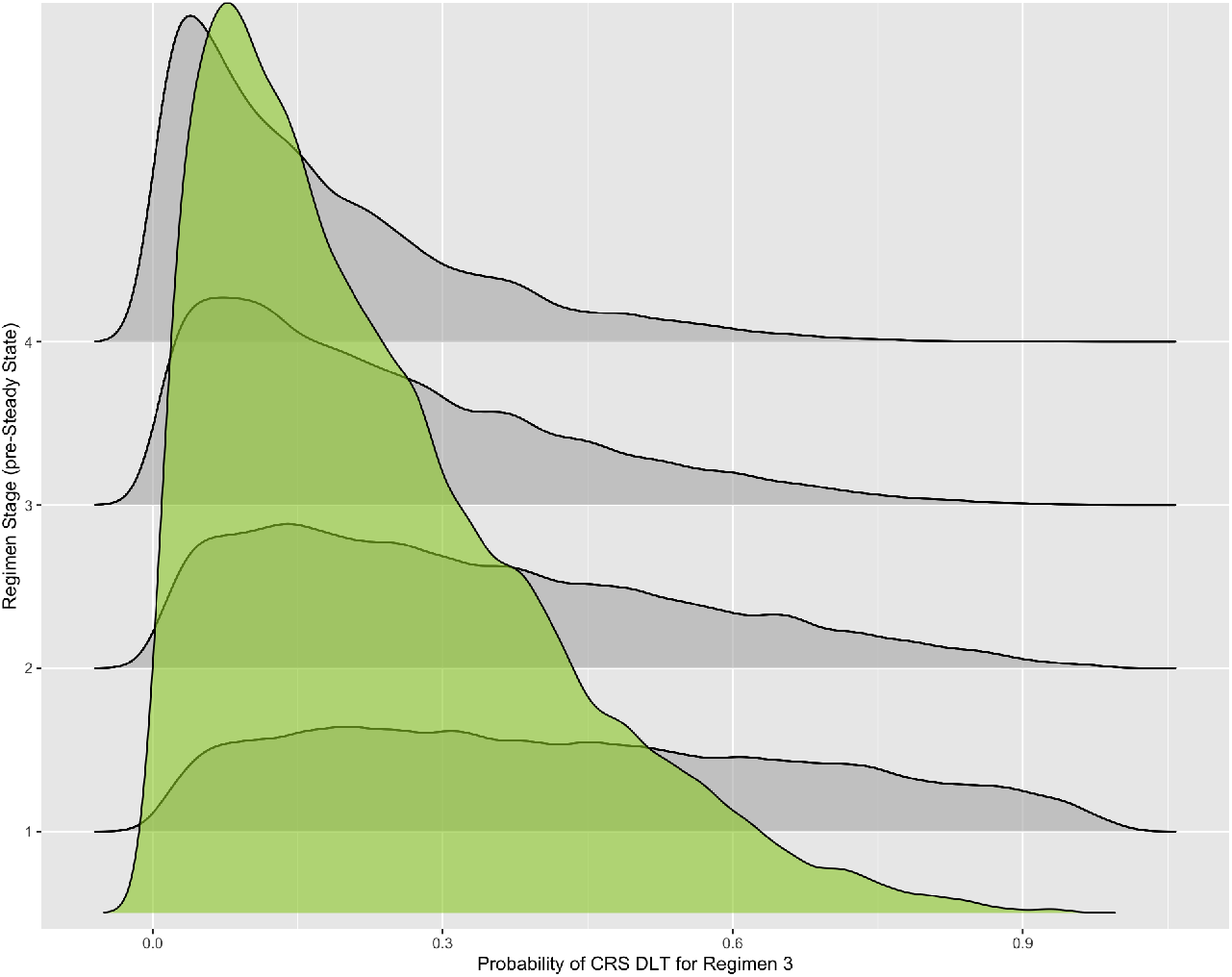
Posterior densities (grey) contributing to overall posterior probability (green) of CRS for DR3.

This level of granularity grants the DEC the ability to interrogate alternative regimens with relative ease. For example, DLTs may have occurred when a regimen with a large increase in relative dose was introduced.

Consider DR5 in Table 1. This regimen includes a relative increase of 700*μ*g at the start of stage 4 (Day 14) - if the cohort on DR4 experience 2 CRS DLTs at stage 4, and no other DLTs have been observed in the study, a reasonable question might be: can we reduce the increase to 500*μ*g? Or, what is the maximum increase at stage 4 which would not lead to overdose?

The modified BLRM has great potential to support this sort of analysis. Given phase I trials rely on well-informed decision making in the face of minimal data, providing the kind of clarity demonstrated here can only be a good thing.

A main weakness of the modified BLRM is the substantial increase in parameters - particularly in a setting where we have very minimal data. This makes prior elicitation harder, and sparse data even sparser (by considering the stage in which a CRS DLT occurs, we are arguably diluting the effect of the DLT on the whole model, compared to a simple BLRM).

However, we think that current approaches are simply not well suited to the problem of modelling a dose regimen in the context of dose-finding. In addition, because the modified BLRM is used flexibly to support decision making, rather than simply codifying a set of rules to follow (such as the 3+3 design, and similar), we think that the advantage of capturing the relationship between dose, prior exposure, and probability of CRS DLT outweighs the problems introduced by a more complex model.

## 5. Conclusion

In conclusion we have presented a modification to the BLRM for use in phase I dose-escalation trials for modern immunotherapies. This novel approach provides all of the functionality of the classic model whilst providing a substantial improvement in granularity, and hence improves the support the model can provide to a dose escalation committee or similar body. Further work is underway to explore better approaches to prior elicitation, and better ways to recommend the optimal next regimen at any point in a trial.

## Data Availability

All simulated data used in analyses are available upon reasonable request to the authors

## Notes

### Competing Interest Statement

The authors have declared no competing interest.

### Funding Statement

Research funded by Cogitars GmbH and its subsidiary company, Cogitars UK Ltd, https://cogitars.com

### Author Declarations

Only simulated data was used

